# Functional difficulty among children and adolescents aged 5–17 years in Ethiopia: Multilevel analysis of the nationally representative 2024–2025 DHS

**DOI:** 10.64898/2026.11.09.26367692

**Authors:** Mahder Dereb, Awoke Mihretu

**Author notes:** Corresponding author: Department of Psychiatry, College of Health Sciences, Zambia Street, Tikur Anbessa Specialised Hospital Campus, Office No. 601 (CHS Building), P.O. Box 9086, Addis Ababa University, Addis Ababa, Ethiopia.

## Abstract

**Background:** Population-based data on functional difficulty among school-age children in Ethiopia are scarce. The 2024–2025 Ethiopia Demographic and Health Survey (EDHS) included, for the first time, the UNICEF/Washington Group Child Functioning Module (CFM) for children aged 5–17 years. We estimated the prevalence and domain distribution of functional difficulty and associated factors in Ethiopia.

**Methods:** We analysed cross-sectional EDHS data from 11,122 children aged 5–17 years (weighted n = 10,726) and 4,739 mothers. Functional difficulty was defined as “a lot of difficulty” or “cannot do at all” in any of 11 domains, or daily anxiety or depression. We estimated design-based prevalence and fitted multilevel logistic regression models (children within mothers within clusters) for any, non-affective and affective difficulty. Interviewer effects were examined with random-intercept models and within-cluster comparisons.

**Results:** Overall, 9.5% (95% CI 8.1–11.1) of children had functional difficulty. Daily depression (6.8%) and anxiety (6.6%) dominated, and 70.3% of affected children had only affective difficulty. Difficulty in at least one non-affective domain affected 2.8% (2.2–3.7), with each domain at 0.5–1.4%. Prevalence ranged from 1.5% in Southwest Regional State, Ethiopia, to 16.5% in Oromia Regional State. In adjusted models, children not attending school had higher odds of non-affective difficulty (AOR 2.82, 95% CI 2.01–3.96), and girls had lower odds (AOR 0.63, 0.48–0.84). Outcomes clustered strongly within families (sibling correlation 67.7%) and clusters (ICC 20.9%). Six of 163 interviewers (7.9% of the weighted sample) contributed 58.2% of weighted cases of daily anxiety or depression; in the same clusters they recorded daily symptoms for 56.4% of children, against 2.1% for other interviewers. Excluding 11 statistically outlying interviewers reduced the prevalence of any difficulty to 4.6% (3.6–5.8) and of daily anxiety or depression to 3.3%. The associations with school non-attendance and sex were unchanged.

**Conclusions:** Functional difficulties across the sensory, physical, cognitive, and affective domains, including anxiety or depression, affect about 2–3% of Ethiopian children aged 5–17 years and are strongly associated with being out of school. Appropriate interventions and resource allocation should be considered.

## Background

Globally, one in ten children has at least one disability. They are more likely than other children to be out of school, to live in poverty, and to lack access to health and social protection [1]. The Convention on the Rights of Persons with Disabilities (RPD) and Sustainable Development Goal (SDG) target 4.5 require countries to identify children with disabilities and monitor their inclusion [1, 2]. Survey questions about diagnosed impairments undercount children in settings where diagnostic services are scarce and are not comparable across countries [3].

UNICEF and the Washington Group on Disability Statistics therefore developed the Child Functioning Module (CFM), which is informed by the biopsychosocial model of disability [3–5]. The module for children aged 5–17 years covers 13 domains: seeing, hearing, walking, self-care, communication, learning, remembering, concentrating, accepting change, controlling behaviour, making friends, and the frequency of anxiety and depression. A child is classified as having a functional difficulty if the respondent reports “a lot of difficulty” or “cannot do at all” in any domain, or if the child shows signs of anxiety or depression daily [4, 5]. The module has undergone cognitive and field testing, including in sub-Saharan Africa [4–6]. Because the affective items ask about the frequency of feelings, they may be more sensitive than the activity items to differences in how questions are translated, understood and administered. Interviewer effects are a recognised source of error in survey measures of sensitive or subjective constructs [7].

Ethiopia ratified the Convention in 2010 and has adopted a Master Plan for Special Needs/Inclusive Education (2016–2025) [8]. National evidence on functional difficulty among school-age children has nevertheless been limited. The 2024–2025 Ethiopia Demographic and Health Survey (EDHS) is the first nationally representative Ethiopian survey to include the CFM for children aged 5–17 years. Its report gives a national prevalence of 9.4%, driven largely by anxiety and depression, with very large regional differences [8]. No study has examined the correlates of functional difficulty in these data while accounting for the clustering of children within families and communities, or assessed how robust the estimates are to measurement error.

This study aimed to (i) estimate the prevalence of functional difficulty among children aged 5–17 years in Ethiopia, overall and by domain; (ii) identify child, maternal, household and community correlates, distinguishing non-affective difficulty from daily anxiety or depression; (iii) quantify variation between families and communities; and (iv) assess interviewer effects and their influence on the estimates.

## Methods

### Data source and design

We used cross-sectional data from the 2024–2025 EDHS, conducted by the Ethiopian Statistical Service with technical support from ICF between August 2024 and April 2025 [8]. The survey used a two-stage stratified cluster design. In the first stage, 805 enumeration areas were selected with probability proportional to size from 27 strata defined by region and urban/rural residence. In the second stage, 28 households were selected in each area. Fieldwork was completed in 797 clusters; eight clusters in Amhara were excluded for security reasons. The household response rate was 98.8%, and 21,395 women aged 15–49 years were interviewed (96.2%) [8]. We followed the STROBE reporting guideline [9]; the completed checklist is provided in Additional file 3.

### Study population and linkage of module records

The CFM was part of the Women’s Questionnaire and administered to a random 50% subsample of households. Mothers completed the module for each biological child aged 5–17 years who lived with them, starting with the youngest [8]. In the women recode file (ETIR8AFL), module responses are stored as up to nine repeated records per woman, each indexed to the birth history (idx97) and the pregnancy history (idx97p). We linked each record to the child via the pregnancy-history index. For 10,450 records (94.0%), this identified a living child aged 5–17 years who lived with the mother. For the remaining 672 records (6.0%), the index pointed to a pregnancy that did not meet these criteria. In every such case, the number of unlinked eligible children matched the number of unlinked records. Among correctly linked records, the order of records matched the children’s age order (youngest first) for 100% of mothers. We therefore assigned the unlinked records to the unlinked eligible children in age order. With this linkage, the weighted numbers of boys (5,512) and girls (5,214), and of children attending (6,466) and not attending school (4,233), exactly reproduced the published figures [8]. The births recode file (ETBR8AFL) contains only one module record per woman, attached via idx97, and that record refers to an eligible child in only 22% of cases. It should therefore not be used for child-level analysis of the CFM. The analytic sample comprised all 11,122 module records (weighted n = 10,726, matching the survey report); 41 children with unknown school attendance were excluded from the regression models (n = 11,081).

### Measurement of functional difficulty

We used the UNICEF and Washington Group on Disability Statistics-developed Child Functioning Module (CFM), developed in consultation with organisations of persons with disabilities and other stakeholder groups. Children who used glasses or hearing aids were asked about seeing or hearing difficulty while using the device; other children were asked about seeing or hearing difficulty directly. Children who used walking equipment or assistance were asked about walking 100 m and 500 m without and with it. Other children were asked to walk 100 m and 500 m, compared with children of the same age. The remaining domains were self-care; communication (being understood by people inside and outside the household); learning; remembering; concentrating; accepting change in routine; controlling behaviour; and making friends. Each had four response options (no difficulty, some difficulty, a lot of difficulty, cannot do at all). Anxiety and depression were assessed by how often the child seemed very anxious, nervous or worried, or very sad or depressed (daily, weekly, monthly, a few times a year, never) [4, 8].

Following UNICEF/Washington Group syntax, a child was considered to have difficulty in a domain if the respondent reported “a lot of difficulty” or “cannot do at all”, or daily anxiety or depression [4, 5]. For device users, responses with the device were used for seeing and hearing, and responses without equipment were used for walking. The primary outcome was functional difficulty in at least one of the 13 domains. Secondary outcomes were (a) difficulty in at least one of the 11 non-affective domains (“non-affective difficulty”), (b) daily anxiety or depression (“affective difficulty”), and (c) difficulty in two or more domains. A broader cut-off (“some difficulty” or worse; anxiety or depression at least weekly) was used descriptively. Our national estimate (9.5%) differs from the published 9.4% only because the survey report did not count difficulty reported by device users while using their device. All domain-specific estimates matched the report to within 0.1 percentage points [8].

### Predictor variables

Predictor variables were selected a priori from the literature on disability and child functioning [1, 10, 11]. Child-level variables were age (5–9, 10–14, 15–17 years), sex and attendance at school during the current school year. We obtained school attendance from the household roster and linked it by the child’s line number. Maternal and household variables were mother’s age (15–29, 30–39, 40–49 years), mother’s highest education, sex of the household head, household size and wealth quintile. Community variables were place of residence and region (14 regions and chartered cities).

### Statistical analysis

Prevalence estimates accounted for the women’s sampling weights, the 792 primary sampling units and the 27 strata (765 design degrees of freedom), with Taylor-linearised standard errors and logit-transformed 95% confidence intervals (CI). Differences across groups were tested with design-adjusted Wald F tests.

Because siblings share a mother and children share communities, we fitted three-level random-intercept logistic regression models (children within mothers within clusters). These were estimated by maximum likelihood with nested Gauss–Hermite quadrature (25 points per level; estimates changed by less than 0.01 on the log-odds scale with 40 points). For each outcome, a null model provided the variance at each level, and a full model included all explanatory variables, with residence and region as fixed effects. Crude odds ratios were obtained from models with one variable at a time. We report adjusted odds ratios (AOR) with 95% CIs. Between-cluster variation is summarised by the intraclass correlation (ICC = σ²(cluster)/[σ²(cluster) + σ²(mother) + π²/3]), the median odds ratio (MOR) and the proportional change in cluster variance (PCV) [12, 13]. The sibling correlation is [σ²(cluster) + σ²(mother)]/[σ²(cluster) + σ²(mother) + π²/3]. Multilevel models were unweighted, an accepted approach when level-specific weights are not available [14]. We fitted design-based single-level logistic regressions, which give population-averaged odds ratios, as a sensitivity analysis. To examine educational exclusion, we modelled non-attendance at school among children aged 7–17 years, with non-affective and affective difficulty as exposures.

Interviewer effects were assessed in three ways. First, we compared the fit (Akaike information criterion, AIC) of two-level models for affective and non-affective difficulty with a random intercept for cluster or for interviewer. Second, for each of the 163 interviewers, we compared the number of children with daily anxiety or depression with the number expected from the other interviewers working in the same clusters, using a Mantel–Haenszel test stratified by cluster; interviewers with Bonferroni-corrected one-sided p < 0.05 were flagged. Because interviewers within a team worked in the same clusters, this comparison holds the community constant. Third, we repeated all analyses after excluding children interviewed by flagged interviewers. Analyses were performed in Python 3.13 (NumPy, SciPy, pandas). The analysis code was written and run with assistance from Claude (Anthropic), an AI model, using the DHS data files obtained by the authors. The authors independently verified all estimates in Stata version 18. The code is provided in Additional file 2.

### Ethical considerations

This study was a secondary analysis of publicly available, de-identified data from the DHS Program, and the researchers did not interact with participants. No separate ethical approval was sought for this analysis; we obtained permission from the DHS Program to access and use the data, and the datasets were downloaded from https://dhsprogram.com/data/available-datasets.cfm. Ethical approval for the original 2024–25 EDHS data collection was granted to the survey implementers by the Ethiopian Public Health Institute Institutional Review Board and the ICF Institutional Review Board, and informed consent was obtained from all respondents at the time of the survey [8].

## Results

### Study population

The 11,122 children (weighted n = 10,726) belonged to 4,739 mothers in 792 clusters. Half (49.9%) were aged 5–9 years, 51.4% were boys, 76.6% lived in rural areas and 60.3% were attending school. Most (61.3%) had mothers with no formal education (Table 1).

**Table 1.** Characteristics of children aged 5–17 years and weighted prevalence of functional difficulty, Ethiopia DHS 2024–2025 (n = 11,122)

| Characteristic | Unweighted n | Weighted % | Any functional difficulty, % (95% CI) | Non-affective difficulty, % (95% CI) | Daily anxiety or depression, % (95% CI) | p-value<sup>a</sup> |
|---|---|---|---|---|---|---|
| **Child characteristics** |  |  |  |  |  |  |
| Age (years) |  |  |  |  |  | 0.040 / 0.254 / 0.097 |
| 5–9 | 5,623 | 49.9 | 10.9 (9.1–13.1) | 3.2 (2.4–4.3) | 9.2 (7.3–11.4) |  |
| 10–14 | 4,238 | 38.3 | 8.4 (6.7–10.5) | 2.6 (1.8–3.7) | 7.0 (5.4–9.1) |  |
| 15–17 | 1,261 | 11.8 | 7.1 (4.9–10.2) | 2.0 (1.1–3.6) | 6.0 (3.9–9.2) |  |
| Sex |  |  |  |  |  | 0.534 / 0.375 / 0.809 |
| Male | 5,723 | 51.4 | 9.9 (8.1–11.9) | 3.1 (2.4–4.0) | 8.1 (6.4–10.2) |  |
| Female | 5,399 | 48.6 | 9.2 (7.5–11.1) | 2.5 (1.6–3.9) | 7.8 (6.3–9.8) |  |
| School attendance (current year) |  |  |  |  |  | 0.297 / 0.288 / 0.061 |
| Attending | 6,747 | 60.3 | 10.0 (8.2–12.2) | 2.5 (1.7–3.8) | 8.8 (7.0–10.9) |  |
| Not attending | 4,334 | 39.5 | 8.8 (7.2–10.7) | 3.3 (2.4–4.4) | 6.8 (5.4–8.6) |  |
| **Maternal and household characteristics** |  |  |  |  |  |  |
| Mother’s age (years) |  |  |  |  |  | 0.004 / 0.024 / 0.027 |
| 15–29 | 1,921 | 17.1 | 14.4 (10.1–20.0) | 4.3 (2.3–7.6) | 11.9 (7.7–17.9) |  |
| 30–39 | 5,896 | 54.8 | 7.3 (5.8–9.0) | 2.0 (1.4–2.7) | 6.2 (4.8–7.9) |  |
| 40–49 | 3,305 | 28.1 | 11.0 (8.1–14.6) | 3.6 (2.4–5.5) | 9.2 (6.5–12.8) |  |
| Mother’s education |  |  |  |  |  | 0.786 / 0.111 / 0.491 |
| No education | 6,766 | 61.3 | 9.1 (7.3–11.4) | 3.0 (2.1–4.3) | 7.8 (6.0–10.0) |  |
| Primary | 3,036 | 27.2 | 10.9 (7.6–15.3) | 2.0 (1.4–2.9) | 9.3 (6.1–13.8) |  |
| Secondary | 777 | 7.4 | 9.1 (6.0–13.5) | 4.8 (2.8–8.1) | 5.7 (3.1–10.1) |  |
| More than secondary | 543 | 4.1 | 7.5 (3.6–14.7) | 2.1 (0.9–5.1) | 6.4 (2.8–13.9) |  |
| Sex of household head |  |  |  |  |  | 0.591 / 0.087 / 0.962 |
| Male | 8,693 | 83.2 | 9.3 (7.8–11.2) | 2.5 (1.9–3.4) | 8.0 (6.4–9.9) |  |
| Female | 2,429 | 16.8 | 10.4 (7.5–14.1) | 4.2 (2.7–6.6) | 7.9 (5.4–11.4) |  |
| Household size (members) |  |  |  |  |  | 0.268 / 0.982 / 0.089 |
| ≤4 | 1,468 | 14.8 | 9.5 (6.9–12.8) | 2.8 (1.7–4.7) | 7.8 (5.4–11.1) |  |
| 5–7 | 5,927 | 55.8 | 10.4 (8.5–12.6) | 2.8 (2.0–3.9) | 9.1 (7.2–11.3) |  |
| ≥8 | 3,727 | 29.4 | 7.9 (5.8–10.8) | 2.9 (1.7–5.0) | 6.0 (4.2–8.7) |  |
| Wealth quintile |  |  |  |  |  | 0.312 / 0.865 / 0.117 |
| Lowest | 4,007 | 23.2 | 9.4 (6.6–13.1) | 3.5 (2.0–6.1) | 7.5 (5.1–10.8) |  |
| Second | 2,222 | 22.6 | 9.2 (6.7–12.4) | 2.4 (1.4–4.1) | 7.8 (5.5–11.1) |  |
| Middle | 1,856 | 20.2 | 11.1 (8.1–15.1) | 2.4 (1.4–4.1) | 9.8 (6.8–13.8) |  |
| Fourth | 1,453 | 19.2 | 10.3 (7.0–14.9) | 2.7 (1.2–6.0) | 9.2 (6.0–14.0) |  |
| Highest | 1,584 | 14.9 | 7.1 (5.1–9.8) | 3.1 (1.9–4.9) | 4.9 (3.0–7.8) |  |
| **Community characteristics** |  |  |  |  |  |  |
| Residence |  |  |  |  |  | 0.705 / 0.268 / 0.545 |
| Urban | 2,718 | 23.4 | 9.0 (6.5–12.3) | 3.7 (2.2–6.2) | 7.2 (4.7–10.8) |  |
| Rural | 8,404 | 76.6 | 9.7 (8.0–11.6) | 2.6 (1.9–3.5) | 8.2 (6.7–10.1) |  |
| Region |  |  |  |  |  | <0.001 / <0.001 / <0.001 |
| Tigray | 922 | 5.8 | 7.1 (5.1–9.9) | 4.6 (3.0–6.8) | 3.3 (2.1–5.2) |  |
| Afar | 951 | 1.1 | 7.7 (5.1–11.5) | 5.5 (3.3–9.0) | 2.6 (1.5–4.5) |  |
| Amhara | 733 | 19.8 | 3.3 (2.1–5.3) | 1.8 (0.9–3.5) | 1.8 (0.9–3.2) |  |
| Oromia | 1,238 | 42.9 | 16.5 (13.5–20.1) | 3.6 (2.3–5.6) | 15.3 (12.3–18.9) |  |
| Somali | 1,222 | 4.6 | 4.3 (2.4–7.6) | 2.6 (1.6–4.3) | 2.2 (1.0–4.8) |  |
| Benishangul-Gumuz | 651 | 1.1 | 6.8 (4.4–10.3) | 2.6 (1.6–4.3) | 4.9 (2.8–8.4) |  |
| Central Ethiopia | 895 | 5.1 | 2.8 (1.7–4.5) | 1.3 (0.7–2.6) | 2.0 (1.2–3.1) |  |
| Sidama | 852 | 4.1 | 3.3 (1.9–5.4) | 1.6 (0.9–2.7) | 2.1 (1.2–3.9) |  |
| South West Ethiopia | 771 | 3.0 | 1.5 (0.9–2.7) | 1.1 (0.5–2.0) | 0.5 (0.1–1.5) |  |
| South Ethiopia | 948 | 8.1 | 4.9 (3.0–7.9) | 1.3 (0.7–2.5) | 3.8 (2.2–6.7) |  |
| Gambella | 590 | 0.4 | 11.4 (8.0–16.0) | 4.0 (2.6–6.1) | 8.3 (5.1–13.3) |  |
| Harari | 493 | 0.3 | 4.2 (2.5–6.8) | 2.1 (1.1–3.9) | 2.5 (1.2–5.4) |  |
| Addis Ababa | 390 | 3.2 | 5.8 (3.1–10.5) | 4.1 (2.1–7.9) | 3.4 (1.5–7.5) |  |
| Dire Dawa | 466 | 0.4 | 5.5 (3.5–8.6) | 3.1 (1.9–5.1) | 2.8 (1.5–4.9) |  |
| Total | 11,122 | 100.0 | 9.5 (8.1–11.1) | 2.8 (2.2–3.7) | 8.0 (6.6–9.6) |  |

Weighted percentages and prevalence account for sampling weights, clustering and stratification. Non-affective difficulty: “a lot of difficulty” or “cannot do at all” in any of 11 domains other than anxiety and depression. School attendance unknown for 41 children.
<sup>a</sup> Design-adjusted Wald F test for any / non-affective / affective difficulty.

### Prevalence and domain distribution

Overall, 9.5% (95% CI 8.1–11.1) of children had functional difficulty in at least one domain (Table 2). Daily depression (6.8%, 5.6–8.3) and daily anxiety (6.6%, 5.4–8.1) were the most common. Each non-affective domain was reported for 0.5% (seeing) to 1.4% (walking, controlling behaviour) of children. In total, 2.8% (2.2–3.7) had difficulty in at least one non-affective domain and 8.0% (6.6–9.6) had daily anxiety or depression. The two affective items largely overlapped: 5.5% of children had both. Consequently, 6.6% had difficulty in two or more domains, and 70.3% (62.0–77.5) of children with any difficulty had affective difficulty only. Daily anxiety or depression was reported for 45.7% of children with non-affective difficulty, compared with 6.9% of other children. With the broader cut-off, 26.4% (23.9–29.0) of children had at least some difficulty, including 12.9% (11.1–14.9) in non-affective domains.

**Table 2.**
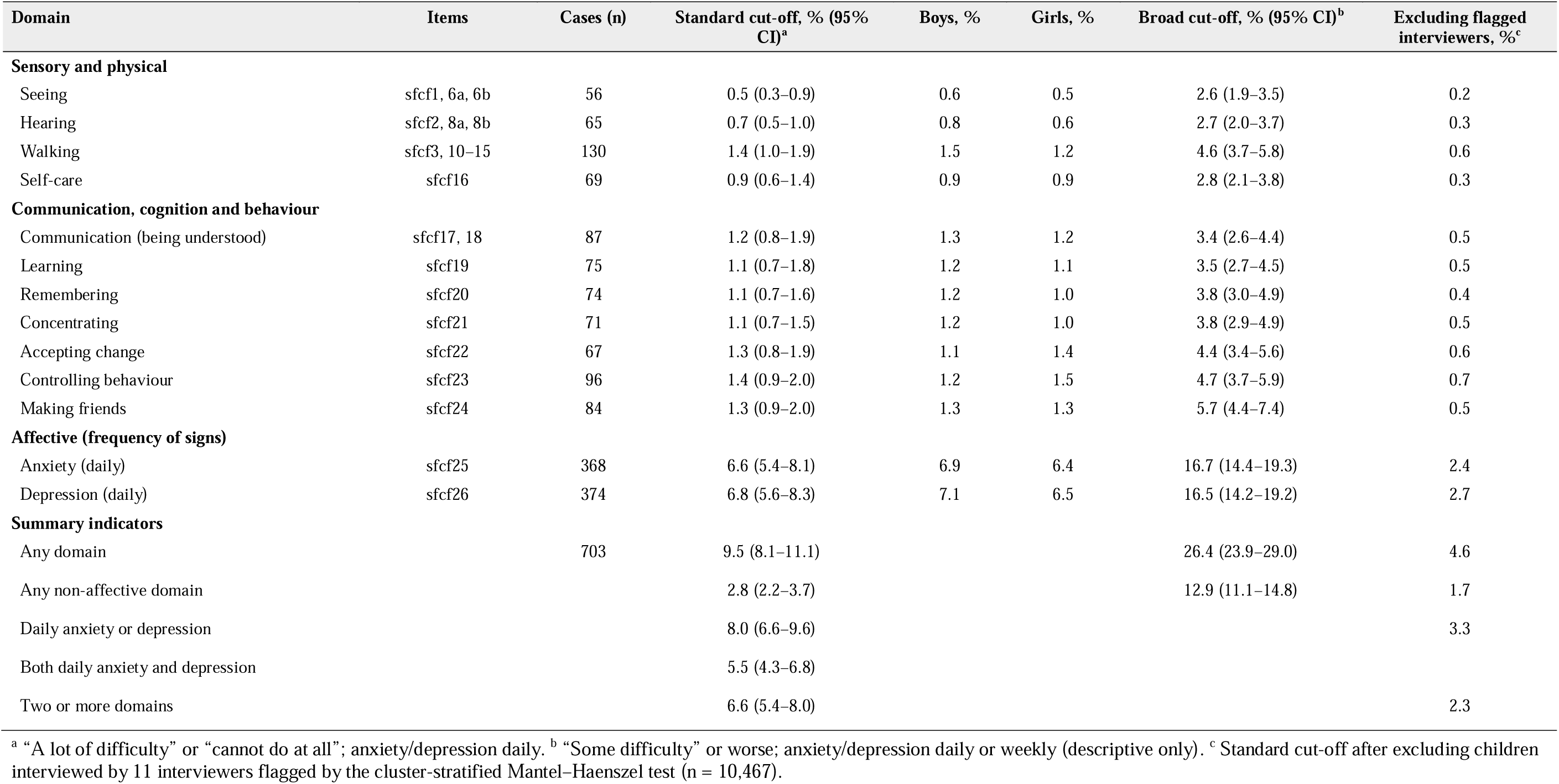
Prevalence of functional difficulty by domain among children aged 5–17 years, Ethiopia DHS 2024–2025

| Domain | Items | Cases (n) | Standard cut-off, % (95% CI)<sup>a</sup> | Boys, % | Girls, % | Broad cut-off, % (95% CI)<sup>b</sup> | Excluding flagged interviewers, %<sup>c</sup> |
|---|---|---|---|---|---|---|---|
| **Sensory and physical** |  |  |  |  |  |  |  |
| Seeing | sfcf1, 6a, 6b | 56 | 0.5 (0.3–0.9) | 0.6 | 0.5 | 2.6 (1.9–3.5) | 0.2 |
| Hearing | sfcf2, 8a, 8b | 65 | 0.7 (0.5–1.0) | 0.8 | 0.6 | 2.7 (2.0–3.7) | 0.3 |
| Walking | sfcf3, 10–15 | 130 | 1.4 (1.0–1.9) | 1.5 | 1.2 | 4.6 (3.7–5.8) | 0.6 |
| Self-care | sfcf16 | 69 | 0.9 (0.6–1.4) | 0.9 | 0.9 | 2.8 (2.1–3.8) | 0.3 |
| **Communication, cognition and behaviour** |  |  |  |  |  |  |  |
| Communication (being understood) | sfcf17, 18 | 87 | 1.2 (0.8–1.9) | 1.3 | 1.2 | 3.4 (2.6–4.4) | 0.5 |
| Learning | sfcf19 | 75 | 1.1 (0.7–1.8) | 1.2 | 1.1 | 3.5 (2.7–4.5) | 0.5 |
| Remembering | sfcf20 | 74 | 1.1 (0.7–1.6) | 1.2 | 1.0 | 3.8 (3.0–4.9) | 0.4 |
| Concentrating | sfcf21 | 71 | 1.1 (0.7–1.5) | 1.2 | 1.0 | 3.8 (2.9–4.9) | 0.5 |
| Accepting change | sfcf22 | 67 | 1.3 (0.8–1.9) | 1.1 | 1.4 | 4.4 (3.4–5.6) | 0.6 |
| Controlling behaviour | sfcf23 | 96 | 1.4 (0.9–2.0) | 1.2 | 1.5 | 4.7 (3.7–5.9) | 0.7 |
| Making friends | sfcf24 | 84 | 1.3 (0.9–2.0) | 1.3 | 1.3 | 5.7 (4.4–7.4) | 0.5 |
| **Affective (frequency of signs)** |  |  |  |  |  |  |  |
| Anxiety (daily) | sfcf25 | 368 | 6.6 (5.4–8.1) | 6.9 | 6.4 | 16.7 (14.4–19.3) | 2.4 |
| Depression (daily) | sfcf26 | 374 | 6.8 (5.6–8.3) | 7.1 | 6.5 | 16.5 (14.2–19.2) | 2.7 |
| **Summary indicators** |  |  |  |  |  |  |  |
| Any domain |  | 703 | 9.5 (8.1–11.1) |  |  | 26.4 (23.9–29.0) | 4.6 |
| Any non-affective domain |  |  | 2.8 (2.2–3.7) |  |  | 12.9 (11.1–14.8) | 1.7 |
| Daily anxiety or depression |  |  | 8.0 (6.6–9.6) |  |  |  | 3.3 |
| Both daily anxiety and depression |  |  | 5.5 (4.3–6.8) |  |  |  |  |
| Two or more domains |  |  | 6.6 (5.4–8.0) |  |  |  | 2.3 |

<sup>a</sup> “A lot of difficulty” or “cannot do at all”; anxiety/depression daily. <sup>b</sup> “Some difficulty” or worse; anxiety/depression daily or weekly (descriptive only). <sup>c</sup> Standard cut-off after excluding children interviewed by 11 interviewers flagged by the cluster-stratified Mantel–Haenszel test (n = 10,467).

### Variation by background characteristics

Prevalence declined with age, from 10.9% at 5–9 years to 7.1% at 15–17 years (p = 0.040). It did not differ significantly by sex (9.9% vs 9.2%), school attendance (10.0% vs 8.8%), mother’s education, wealth or residence (Table 1). Prevalence was higher among children of mothers aged 15–29 (14.4%) or 40–49 years (11.0%) than among those of mothers aged 30–39 years (7.3%; p = 0.004). Regional differences were large (p < 0.001), from 1.5% in South West Ethiopia, 2.8% in Central Ethiopia and 3.3% in Amhara and Sidama to 11.4% in Gambella and 16.5% in Oromia. In Oromia, 15.3% of children had daily anxiety or depression, whereas non-affective difficulty (3.6%) was similar to that in other regions.

### Multilevel analysis

Functional difficulty was strongly clustered (Table 3). In the null model for any difficulty, the variance was 2.12 between clusters and 4.77 between mothers. This gave a cluster ICC of 20.9%, a sibling correlation of 67.7% and a cluster MOR of 4.02. Clustering was strongest for affective difficulty (cluster ICC 27.8%; sibling correlation 77.3%) and weaker for non-affective difficulty (14.9%; 54.5%). The explanatory variables explained 52.2% of the cluster-level variance for any difficulty and 61.8% for affective difficulty, mainly through region, but only 22.4% for non-affective difficulty.

**Table 3.** Factors associated with functional difficulty among children aged 5–17 years: three-level mixed-effects logistic regression (children within mothers within clusters), Ethiopia DHS 2024–2025 (n = 11,081)

| Variable | Any difficulty, COR (95% CI) | Any difficulty, AOR (95% CI) | Non-affective difficulty, AOR (95% CI) | Daily anxiety or depression, AOR (95% CI) |
|---|---|---|---|---|
| **Child level** |  |  |  |  |
| Age (years) |  |  |  |  |
| 5–9 (ref) | 1.00 | 1.00 | 1.00 | 1.00 |
| 10–14 | 0.84 (0.67–1.05) | 0.99 (0.78–1.26) | 0.90 (0.66–1.24) | 1.00 (0.73–1.35) |
| 15–17 | 0.67 (0.46–0.96)* | 0.70 (0.48–1.02) | 0.66 (0.40–1.09) | 0.73 (0.45–1.17) |
| Sex |  |  |  |  |
| Male (ref) | 1.00 | 1.00 | 1.00 | 1.00 |
| Female | 0.87 (0.70–1.08) | 0.86 (0.70–1.07) | 0.63 (0.48–0.84)** | 1.09 (0.82–1.44) |
| School attendance (current year) |  |  |  |  |
| Attending (ref) | 1.00 | 1.00 | 1.00 | 1.00 |
| Not attending | 1.72 (1.33–2.22)*** | 1.91 (1.46–2.50)*** | 2.82 (2.01–3.96)*** | 1.28 (0.90–1.82) |
| **Maternal and household level** |  |  |  |  |
| Mother’s age (years) |  |  |  |  |
| 15–29 (ref) | 1.00 | 1.00 | 1.00 | 1.00 |
| 30–39 | 0.59 (0.41–0.85)** | 0.78 (0.53–1.13) | 0.77 (0.50–1.19) | 1.00 (0.60–1.68) |
| 40–49 | 0.89 (0.60–1.31) | 1.37 (0.89–2.11) | 1.34 (0.82–2.19) | 1.84 (1.02–3.32)* |
| Mother’s education |  |  |  |  |
| No education (ref) | 1.00 | 1.00 | 1.00 | 1.00 |
| Primary | 1.32 (0.95–1.85) | 1.57 (1.09–2.24)* | 1.29 (0.85–1.96) | 1.34 (0.83–2.17) |
| Secondary | 1.30 (0.76–2.23) | 1.40 (0.77–2.55) | 1.69 (0.86–3.31) | 1.09 (0.49–2.43) |
| More than secondary | 1.03 (0.53–1.98) | 1.35 (0.65–2.80) | 1.68 (0.73–3.86) | 0.90 (0.34–2.40) |
| Sex of household head |  |  |  |  |
| Male (ref) | 1.00 | 1.00 | 1.00 | 1.00 |
| Female | 1.47 (1.05–2.06)* | 1.28 (0.90–1.83) | 1.30 (0.87–1.93) | 1.14 (0.71–1.85) |
| Household size (members) |  |  |  |  |
| ≤4 (ref) | 1.00 | 1.00 | 1.00 | 1.00 |
| 5–7 | 0.65 (0.44–0.94)* | 0.68 (0.46–1.01) | 0.85 (0.54–1.36) | 0.58 (0.35–0.97)* |
| ≥8 | 0.67 (0.43–1.03) | 0.73 (0.46–1.16) | 0.88 (0.51–1.51) | 0.58 (0.32–1.07) |
| Wealth quintile |  |  |  |  |
| Lowest (ref) | 1.00 | 1.00 | 1.00 | 1.00 |
| Second | 0.82 (0.53–1.26) | 0.97 (0.61–1.53) | 1.01 (0.60–1.69) | 0.90 (0.49–1.66) |
| Middle | 1.01 (0.63–1.61) | 1.25 (0.77–2.03) | 1.23 (0.70–2.16) | 1.25 (0.66–2.38) |
| Fourth | 0.89 (0.54–1.49) | 0.92 (0.52–1.62) | 1.11 (0.58–2.12) | 0.76 (0.36–1.61) |
| Highest | 0.96 (0.59–1.56) | 0.91 (0.43–1.90) | 0.95 (0.41–2.23) | 0.74 (0.28–1.93) |
| **Community level** |  |  |  |  |
| Residence |  |  |  |  |
| Urban (ref) | 1.00 | 1.00 | 1.00 | 1.00 |
| Rural | 0.82 (0.56–1.22) | 0.66 (0.38–1.13) | 0.77 (0.42–1.40) | 0.49 (0.24–1.00)* |
| Region |  |  |  |  |
| Tigray | 1.97 (0.78–4.95) | 2.53 (0.94–6.87) | 1.57 (0.56–4.40) | 2.47 (0.61–9.93) |
| Afar | 2.43 (0.95–6.20) | 3.21 (1.12–9.22)* | 1.92 (0.65–5.71) | 2.47 (0.56–10.93) |
| Amhara | 0.61 (0.22–1.69) | 0.71 (0.24–2.13) | 0.34 (0.10–1.13) | 1.10 (0.24–5.03) |
| Oromia | 8.53 (3.54–20.58)*** | 12.58 (4.72–33.56)*** | 1.17 (0.41–3.33) | 46.52 (11.88–182.18)*** |
| Somali | 0.80 (0.31–2.09) | 0.94 (0.32–2.78) | 0.57 (0.19–1.77) | 1.00 (0.22–4.59) |
| Benishangul-Gumuz | 1.62 (0.60–4.42) | 2.19 (0.74–6.46) | 1.01 (0.32–3.17) | 3.42 (0.78–15.03) |
| Central Ethiopia | 0.50 (0.18–1.40) | 0.65 (0.22–1.95) | 0.36 (0.11–1.18) | 1.50 (0.35–6.51) |
| Sidama | 0.61 (0.23–1.67) | 0.76 (0.26–2.25) | 0.43 (0.13–1.39) | 1.20 (0.27–5.29) |
| South West Ethiopia | 0.24 (0.08–0.75)* | 0.30 (0.09–1.03) | 0.26 (0.07–0.94)* | 0.15 (0.02–1.06) |
| South Ethiopia | 0.95 (0.36–2.50) | 1.15 (0.40–3.31) | 0.34 (0.10–1.10) | 2.48 (0.60–10.31) |
| Gambella | 3.56 (1.35–9.43)* | 4.57 (1.59–13.15)** | 1.65 (0.54–5.01) | 7.43 (1.74–31.64)** |
| Harari | 0.80 (0.26–2.45) | 1.05 (0.33–3.39) | 0.53 (0.15–1.90) | 1.70 (0.35–8.35) |
| Addis Ababa (ref) | 1.00 | 1.00 | 1.00 | 1.00 |
| Dire Dawa | 1.37 (0.47–4.03) | 1.89 (0.63–5.72) | 1.07 (0.34–3.43) | 2.48 (0.54–11.42) |
| **Random effects (null model → full model)** |  |  |  |  |
| Cluster variance |  | 2.12 → 1.01 | 1.08 → 0.84 | 4.03 → 1.54 |
| Mother variance |  | 4.77 → 4.71 | 2.87 → 3.08 | 7.17 → 6.86 |
| ICC, cluster (%) |  | 20.9 → 11.3 | 14.9 → 11.6 | 27.8 → 13.1 |
| Sibling correlation, ICC mother + cluster (%) |  | 67.7 → 63.5 | 54.5 → 54.4 | 77.3 → 71.8 |
| MOR, cluster |  | 4.02 → 2.61 | 2.69 → 2.39 | 6.78 → 3.26 |
| AIC |  | 4625 → 4495 | 2684 → 2625 | 3246 → 3138 |
| PCV, cluster (%) |  | 52.2 | 22.3 | 61.8 |

COR: crude odds ratio (three-level model with the variable alone); AOR: adjusted odds ratio (all variables shown); ref: reference category; ICC: intraclass correlation coefficient; MOR: median odds ratio; PCV: proportional change in cluster variance from null to full model; AIC: Akaike information criterion. <sup>\*</sup>p < 0.05; <sup>\*\*</sup>p < 0.01; <sup>\*\*\*</sup>p < 0.001.

In the full models, children not attending school had higher odds of any difficulty (AOR 1.91, 95% CI 1.46–2.50) and of non-affective difficulty (AOR 2.82, 2.01–3.96). Girls had lower odds of non-affective difficulty than boys (AOR 0.63, 0.48–0.84). Children of mothers with primary education had higher odds of any difficulty than those of mothers with no education (AOR 1.57, 1.09–2.24). For affective difficulty, odds were higher among children of mothers aged 40–49 years (AOR 1.84, 1.02–3.32) and lower in households of 5–7 members (AOR 0.58, 0.35–0.97) and in rural areas (AOR 0.49, 0.24–1.00). Compared with Addis Ababa, odds of any difficulty were higher in Oromia (AOR 12.58, 4.72–33.56), Gambella (AOR 4.57, 1.59–13.15) and Afar (AOR 3.21, 1.12–9.22). The excess in Oromia was confined to affective difficulty (AOR 46.52 vs 1.17 for non-affective difficulty). Wealth, sex of household head and age were not independently associated with any outcome. Population-averaged odds ratios from design-based models were smaller, as expected given the large random-effect variances (Additional file 1, Table S2).

Among children aged 7–17 years (n = 8,665), non-attendance at school was strongly associated with non-affective difficulty (AOR 4.21, 2.61–6.81) but not with affective difficulty (AOR 1.08, 0.71–1.66) (Additional file 1, Table S1). Unadjusted non-attendance was similar among children with non-affective difficulty (34.9%) and without any difficulty (33.6%), because non-affective difficulty was reported more often in better-off, more educated and urban households where attendance is high.

### Interviewer effects

For affective difficulty, a model with an interviewer random intercept fitted the data much better than one with a cluster random intercept (AIC 2,947 vs 3,401). The interviewer ICC was 43.9%, compared with 24.9% for non-affective difficulty (Table 4). The proportion of interviewed children with daily anxiety or depression ranged from 0% to 72.6% across the 16 interviewers in Oromia. Six of 163 interviewers nationally (four in Oromia, one each in Somali and Harari) recorded daily anxiety or depression for more than 30% of the children they interviewed. These six interviewers covered 273 children (7.9% of the weighted sample) but accounted for 58.2% of the weighted cases of daily anxiety or depression. In the 54 clusters where they worked, 56.4% of the children they interviewed had daily anxiety or depression, compared with 2.1% of the children interviewed by other interviewers in the same clusters. They also recorded non-affective difficulty for 13.2% of children, compared with 1.5%. The cluster-stratified Mantel–Haenszel test flagged 11 interviewers (8 regions; 655 children, 11.1% of the weighted sample; Additional file 1, Table S3).

**Table 4.** Interviewer effects and sensitivity of prevalence estimates, Ethiopia DHS 2024–2025

| **Indicator** | **All children** | **Excluding flagged interviewers<sup>a</sup>** |
|---|---|---|
| **Prevalence, % (95% CI)** |  |  |
| Any functional difficulty | 9.5 (8.1–11.1) | 4.6 (3.6–5.8) |
| Non-affective difficulty | 2.8 (2.2–3.7) | 1.7 (1.3–2.2) |
| Daily anxiety or depression | 8.0 (6.6–9.6) | 3.3 (2.4–4.5) |
| Two or more domains | 6.6 (5.4–8.1) | 2.3 (1.7–3.2) |
| Any difficulty, Oromia | 16.5 (13.5–20.1) | 6.4 |
| Any difficulty, range across regions | 1.5–16.5 | 1.5–9.4 |
| **Variance components, daily anxiety or depression (full covariates)** |  |  |
| Two-level model, cluster intercept: ICC (%) / AIC | 29.3 / 3,401 | — |
| Two-level model, interviewer intercept: ICC (%) / AIC | 43.9 / 2,947 | — |
| Non-affective difficulty, interviewer intercept: ICC (%) | 24.9 | — |
| **Six interviewers recording daily anxiety or depression for >30% of children** |  |  |
| Children interviewed (% of weighted sample) | 273 (7.9) | — |
| Share of weighted cases of daily anxiety or depression (%) | 58.2 | — |
| Daily anxiety or depression in their 54 clusters: their children vs other interviewers' children (%) | 56.4 vs 2.1 | — |
| Non-affective difficulty in the same comparison (%) | 13.2 vs 1.5 | — |

<sup>a</sup> 11 of 163 interviewers with Bonferroni-corrected one-sided p < 0.05 in a Mantel–Haenszel test comparing their cases of daily anxiety or depression with those of other interviewers in the same clusters; 655 children (11.1% of the weighted sample) excluded.

After excluding children interviewed by flagged interviewers, the prevalence of any difficulty was 4.6% (3.6–5.8), of non-affective difficulty 1.7% (1.3–2.2), and of daily anxiety or depression 3.3% (2.4–4.5) (Tables 2 and 4). In Oromia, prevalence fell from 16.5% to 6.4%, and the range across regions narrowed to 1.5–9.4%. The associations of non-attendance with any difficulty (AOR 1.84, 1.39–2.43) and with non-affective difficulty (AOR 2.96, 2.07–4.24), and of female sex with non-affective difficulty (AOR 0.61, 0.46–0.83), were essentially unchanged. The Oromia excess in affective difficulty was much smaller (AOR 6.16, 1.79–21.13) (Additional file 1, Table S1).

## Discussion

We found that 1 in 10 children aged 5–17 years (9.5%) had functional difficulties in Ethiopia, per the UNICEF/Washington Group CFM definition. This is close to the global estimate [1]. About 2.8% of children had difficulty in sensory, physical, communication, cognitive or behavioural domains, and 70% of children classified as having a functional difficulty had affective difficulties, including anxiety and depression, alone. We also found that functional difficulties were strongly clustered within families and communities. Anxiety and depression items, and to a lesser extent the other items, showed large interviewer effects that account for much of the national prevalence and most regional differences.

The interviewer findings are the most consequential. Daily anxiety or depression was recorded for over half of the children seen by a few interviewers and for about 2% of the children seen by their colleagues in the same clusters, where children shared communities, services and exposure to adversity. Differences in true prevalence cannot plausibly explain this pattern, because household allocation within clusters is not related to child characteristics. The pattern points instead to differences in how the questions were asked, translated or recorded, a known risk for subjective items [7]. Possible mechanisms include reading the frequency options incompletely, translating “very anxious” or “very sad” as milder states, or recording default answers. Other evidence supports this interpretation. Siblings were highly correlated (sibling correlation 77% for affective difficulty), as expected if a single respondent–interviewer pair answers for all children in a family. The regional excess was confined to affective items. Prevalence in conflict-affected Tigray and Amhara was low, which argues against an explanation based only on exposure to adversity. Excluding flagged interviewers halved the national prevalence and removed most of the regional variation. We do not regard the trimmed estimates as definitive, because exclusion may also remove some true cases. They show, however, that published CFM estimates for Ethiopia, especially for anxiety, depression and regional comparisons, carry substantial measurement uncertainty that is not captured by sampling errors.

The non-affective estimates were more robust, and their correlates were consistent across analyses. Children with sensory, physical or cognitive difficulty had about three to four times the odds of being out of school, after adjustment and after excluding flagged interviewers. This agrees with evidence from 30 countries that disability is among the strongest predictors of school exclusion [11]. It also underlines the importance of Ethiopia’s inclusive education master plan [8]. The raw comparison showed no attendance gap, because non-affective difficulty was reported more often in urban, wealthier and more educated households where attendance is high. This reversal of the expected poverty–disability gradient [10] most likely reflects differences in awareness and reporting: caregivers with more education and access to services may recognise and report difficulties more readily. Higher mortality among children with severe impairments in poorer settings may also contribute. Boys had higher odds of non-affective difficulty than girls, in line with the male excess in developmental and behavioural conditions reported elsewhere [1].

The predominance of affective items has practical implications. If CFM estimates are reported only as a single indicator, they may be read as describing physical or sensory disability when they largely reflect affective symptoms. These symptoms were also the most error-prone component. We recommend that the EDHS and future surveys report non-affective and affective difficulty separately. Interviewer training on the affective items should be strengthened and supported by case vignette, and interviewer-level monitoring should be included in field supervision. The anxiety and depression items should be validated cognitively and against clinical measures in Ethiopia’s main languages. Child and adolescent mental health needs in Ethiopia are real and largely unmet [15, 16], but planning services for them requires better measurement than these data currently provide.

### Strengths and limitations

Strengths include nationally representative data, an internationally standardised instrument, a linkage of module records to children that reproduced the published totals exactly, three-level models that separate family and community variation, and an explicit assessment of interviewer effects. Limitations include the cross-sectional design, and reliance on maternal report with no clinical assessment. Children who did not live with their mothers, including some of those most likely to have disabilities, were excluded. The cross-language validity of the items is unknown. We could not fit models with crossed cluster and interviewer effects, so interviewer and community influences are only partly separated, although the within-cluster comparison addresses this directly. Flagging interviewers relies on a statistical criterion and may misclassify some interviewers who worked with genuinely high-need families. Finally, 6% of module records required probabilistic linkage, although the rule we applied was supported by the perfect concordance between record order and age among correctly linked records.

## Conclusions

About 3% of Ethiopian children aged 5–17 years have difficulty in sensory, physical, communication, cognitive or behavioural functioning, and these children are much more likely to be out of school. Headline estimates of about 10% are driven by reports of daily anxiety and depression, which were highly sensitive to the interviewer. Policy and research should treat the non-affective and affective components separately, strengthen inclusive education for children with functional difficulty, and invest in better measurement of child mental health in national surveys.

## Abbreviations

AIC: Akaike information criterion;
AOR: adjusted odds ratio;
CFM: Child Functioning Module;
CI: confidence interval;
EDHS: Ethiopia Demographic and Health Survey;
ICC: intraclass correlation coefficient;
MOR: median odds ratio;
PCV: proportional change in variance;
UNICEF: United Nations Children’s Fund.

## Declarations

### Ethics approval and consent to participate

Not applicable for this secondary analysis. The study used publicly available, de-identified data from the DHS Program, with no interaction between the researchers and the participants; permission to use the data was obtained from the DHS Program. The original 2024–25 EDHS data collection was approved by the Ethiopian Public Health Institute Institutional Review Board and the ICF Institutional Review Board, and informed consent was obtained from all participants by the survey implementers [8].

### Responsible AI use

Claude (Anthropic) assisted with data management and analysis code, and with drafting the manuscript. The authors reviewed and verified all analyses, results and text, and take full responsibility for them.

### Consent for publication

Not applicable.

### Availability of data and materials

The datasets are available from the DHS Program (https://dhsprogram.com) on registration. Analysis code, including the record-linkage procedure, is provided in Additional file 2.

## Funding

None

### Authors’ contributions

MD conceptualised the study and led the data analysis. AM drafted the manuscript. Both authors read and approved the final manuscript.

## Acknowledgements

We thank the DHS Program and the Ethiopian Statistical Service for access to the data, and the families who took part in the survey.

---

**Additional file 1, Table S1.**
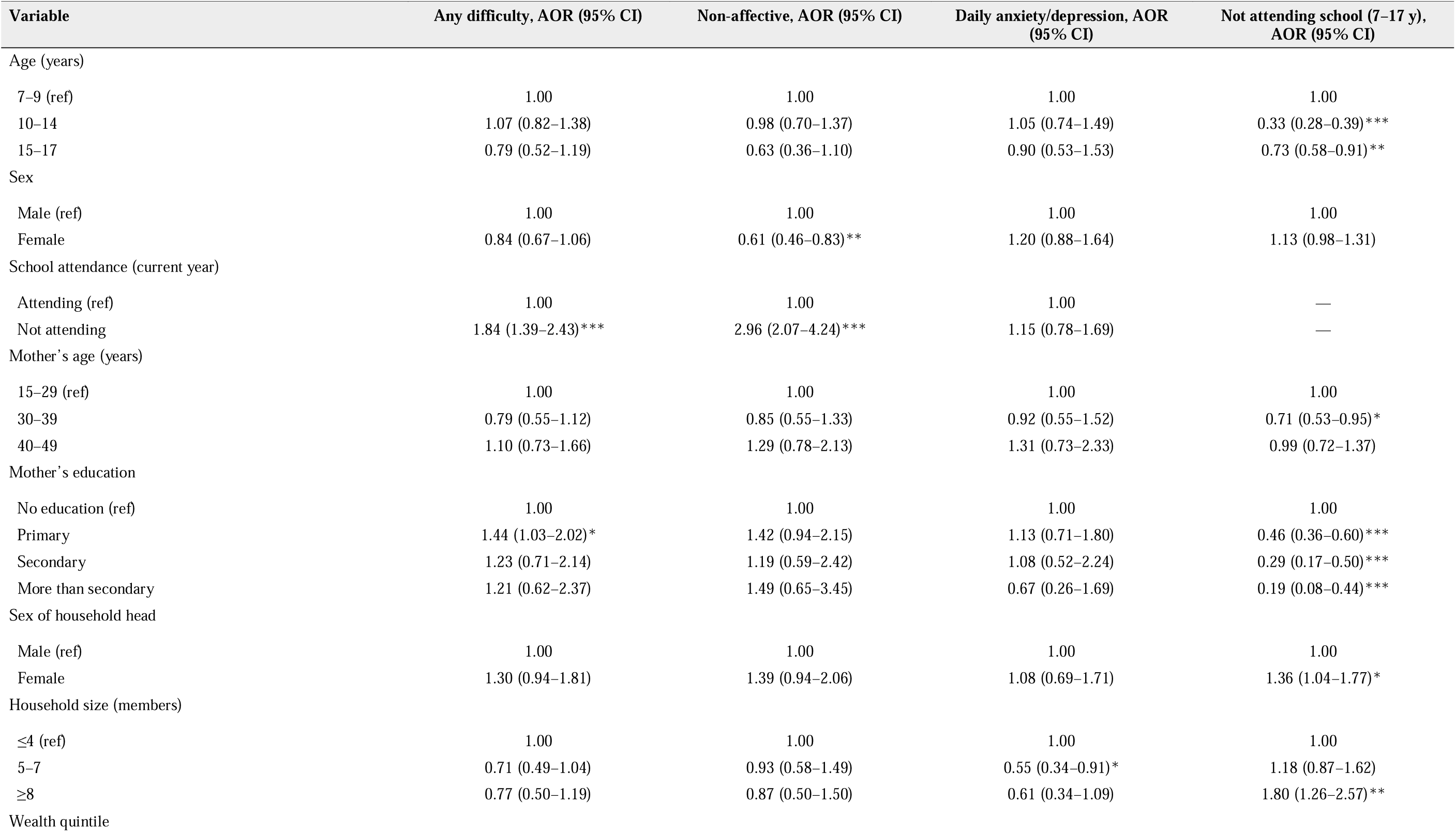
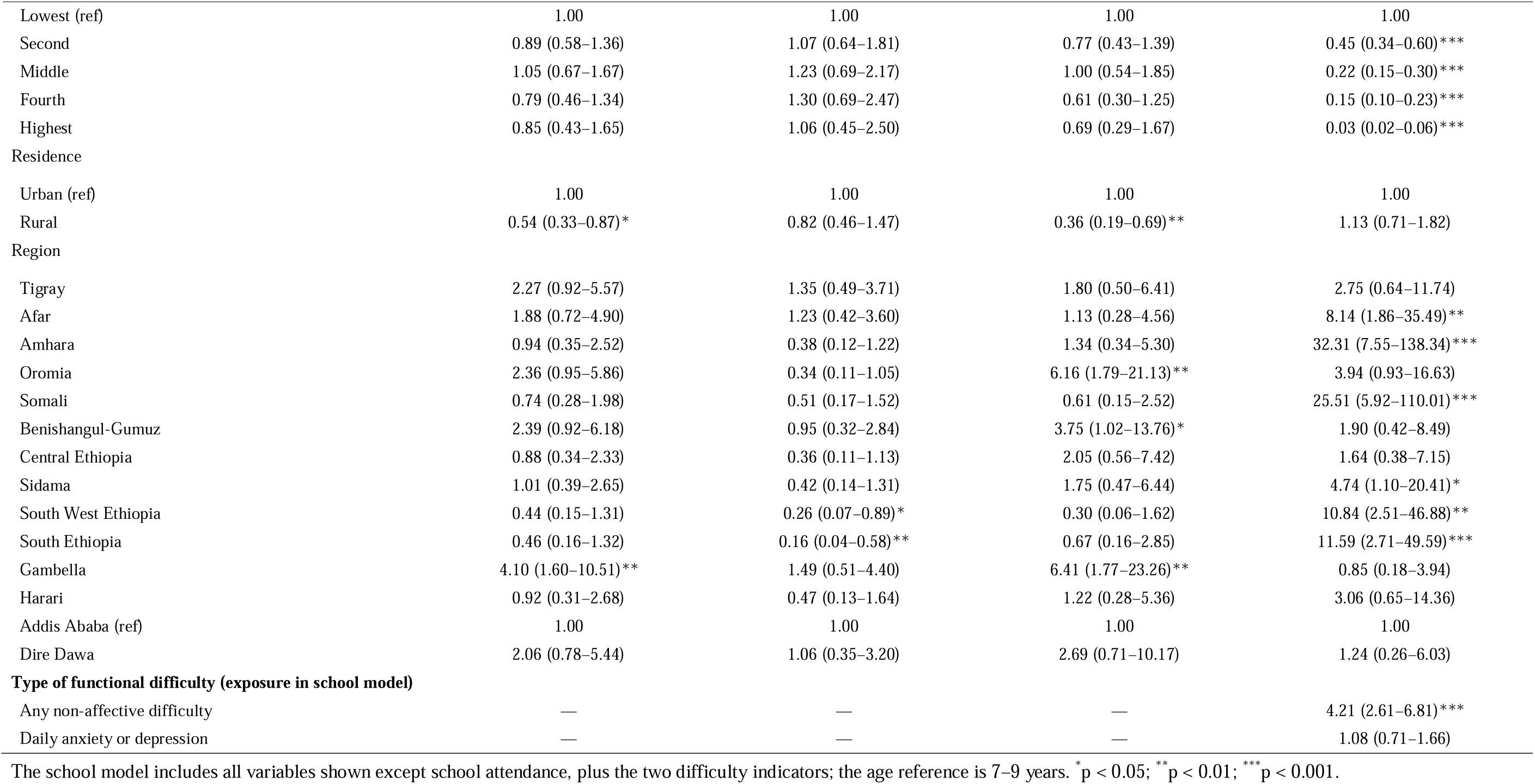
Sensitivity analysis excluding flagged interviewers (three-level models, n = 10,428) and model for school non-attendance among children aged 7–17 years (all interviewers, n = 8,665)

| Variable | Any difficulty, AOR (95% CI) | Non-affective, AOR (95% CI) | Daily anxiety/depression, AOR (95% CI) | Not attending school (7–17 y), AOR (95% CI) |
|---|---|---|---|---|
| Age (years) |  |  |  |  |
| 7–9 (ref) | 1.00 | 1.00 | 1.00 | 1.00 |
| 10–14 | 1.07 (0.82–1.38) | 0.98 (0.70–1.37) | 1.05 (0.74–1.49) | 0.33 (0.28–0.39)*** |
| 15–17 | 0.79 (0.52–1.19) | 0.63 (0.36–1.10) | 0.90 (0.53–1.53) | 0.73 (0.58–0.91)** |
| Sex |  |  |  |  |
| Male (ref) | 1.00 | 1.00 | 1.00 | 1.00 |
| Female | 0.84 (0.67–1.06) | 0.61 (0.46–0.83)** | 1.20 (0.88–1.64) | 1.13 (0.98–1.31) |
| School attendance (current year) |  |  |  |  |
| Attending (ref) | 1.00 | 1.00 | 1.00 | — |
| Not attending | 1.84 (1.39–2.43)*** | 2.96 (2.07–4.24)*** | 1.15 (0.78–1.69) | — |
| Mother’s age (years) |  |  |  |  |
| 15–29 (ref) | 1.00 | 1.00 | 1.00 | 1.00 |
| 30–39 | 0.79 (0.55–1.12) | 0.85 (0.55–1.33) | 0.92 (0.55–1.52) | 0.71 (0.53–0.95)* |
| 40–49 | 1.10 (0.73–1.66) | 1.29 (0.78–2.13) | 1.31 (0.73–2.33) | 0.99 (0.72–1.37) |
| Mother’s education |  |  |  |  |
| No education (ref) | 1.00 | 1.00 | 1.00 | 1.00 |
| Primary | 1.44 (1.03–2.02)* | 1.42 (0.94–2.15) | 1.13 (0.71–1.80) | 0.46 (0.36–0.60)*** |
| Secondary | 1.23 (0.71–2.14) | 1.19 (0.59–2.42) | 1.08 (0.52–2.24) | 0.29 (0.17–0.50)*** |
| More than secondary | 1.21 (0.62–2.37) | 1.49 (0.65–3.45) | 0.67 (0.26–1.69) | 0.19 (0.08–0.44)*** |
| Sex of household head |  |  |  |  |
| Male (ref) | 1.00 | 1.00 | 1.00 | 1.00 |
| Female | 1.30 (0.94–1.81) | 1.39 (0.94–2.06) | 1.08 (0.69–1.71) | 1.36 (1.04–1.77)* |
| Household size (members) |  |  |  |  |
| ≤4 (ref) | 1.00 | 1.00 | 1.00 | 1.00 |
| 5–7 | 0.71 (0.49–1.04) | 0.93 (0.58–1.49) | 0.55 (0.34–0.91)* | 1.18 (0.87–1.62) |
| ≥8 | 0.77 (0.50–1.19) | 0.87 (0.50–1.50) | 0.61 (0.34–1.09) | 1.80 (1.26–2.57)** |
| Wealth quintile |  |  |  |  |
| Lowest (ref) | 1.00 | 1.00 | 1.00 | 1.00 |
| Second | 0.89 (0.58–1.36) | 1.07 (0.64–1.81) | 0.77 (0.43–1.39) | 0.45 (0.34–0.60)*** |
| Middle | 1.05 (0.67–1.67) | 1.23 (0.69–2.17) | 1.00 (0.54–1.85) | 0.22 (0.15–0.30)*** |
| Fourth | 0.79 (0.46–1.34) | 1.30 (0.69–2.47) | 0.61 (0.30–1.25) | 0.15 (0.10–0.23)*** |
| Highest | 0.85 (0.43–1.65) | 1.06 (0.45–2.50) | 0.69 (0.29–1.67) | 0.03 (0.02–0.06)*** |
| Residence |  |  |  |  |
| Urban (ref) | 1.00 | 1.00 | 1.00 | 1.00 |
| Rural | 0.54 (0.33–0.87)* | 0.82 (0.46–1.47) | 0.36 (0.19–0.69)** | 1.13 (0.71–1.82) |
| Region |  |  |  |  |
| Tigray | 2.27 (0.92–5.57) | 1.35 (0.49–3.71) | 1.80 (0.50–6.41) | 2.75 (0.64–11.74) |
| Afar | 1.88 (0.72–4.90) | 1.23 (0.42–3.60) | 1.13 (0.28–4.56) | 8.14 (1.86–35.49)** |
| Amhara | 0.94 (0.35–2.52) | 0.38 (0.12–1.22) | 1.34 (0.34–5.30) | 32.31 (7.55–138.34)*** |
| Oromia | 2.36 (0.95–5.86) | 0.34 (0.11–1.05) | 6.16 (1.79–21.13)** | 3.94 (0.93–16.63) |
| Somali | 0.74 (0.28–1.98) | 0.51 (0.17–1.52) | 0.61 (0.15–2.52) | 25.51 (5.92–110.01)*** |
| Benishangul-Gumuz | 2.39 (0.92–6.18) | 0.95 (0.32–2.84) | 3.75 (1.02–13.76)* | 1.90 (0.42–8.49) |
| Central Ethiopia | 0.88 (0.34–2.33) | 0.36 (0.11–1.13) | 2.05 (0.56–7.42) | 1.64 (0.38–7.15) |
| Sidama | 1.01 (0.39–2.65) | 0.42 (0.14–1.31) | 1.75 (0.47–6.44) | 4.74 (1.10–20.41)* |
| South West Ethiopia | 0.44 (0.15–1.31) | 0.26 (0.07–0.89)* | 0.30 (0.06–1.62) | 10.84 (2.51–46.88)** |
| South Ethiopia | 0.46 (0.16–1.32) | 0.16 (0.04–0.58)** | 0.67 (0.16–2.85) | 11.59 (2.71–49.59)*** |
| Gambella | 4.10 (1.60–10.51)** | 1.49 (0.51–4.40) | 6.41 (1.77–23.26)** | 0.85 (0.18–3.94) |
| Harari | 0.92 (0.31–2.68) | 0.47 (0.13–1.64) | 1.22 (0.28–5.36) | 3.06 (0.65–14.36) |
| Addis Ababa (ref) | 1.00 | 1.00 | 1.00 | 1.00 |
| Dire Dawa | 2.06 (0.78–5.44) | 1.06 (0.35–3.20) | 2.69 (0.71–10.17) | 1.24 (0.26–6.03) |
| **Type of functional difficulty (exposure in school model)** |  |  |  |  |
| Any non-affective difficulty | — | — | — | 4.21 (2.61–6.81)*** |
| Daily anxiety or depression | — | — | — | 1.08 (0.71–1.66) |

The school model includes all variables shown except school attendance, plus the two difficulty indicators; the age reference is 7–9 years. <sup>\*</sup>p < 0.05; <sup>\*\*</sup>p < 0.01; <sup>\*\*\*</sup>p < 0.001.

**Additional file 1, Table S2.** Population-averaged odds ratios from design-based single-level logistic regression (selected variables; models include all covariates Table 3)

| Variable | Any difficulty | Non-affective | Daily anxiety/depression | Any, excl. flagged | Non-affective, excl. flagged |
|---|---|---|---|---|---|
| Not attending school (vs attending) | 0.95 (0.72–1.24) | 1.40 (0.80–2.46) | 0.82 (0.62–1.08) | 1.42 (1.01–1.99)* | 2.62 (1.44–4.76)** |
| Female (vs male) | 0.93 (0.71–1.21) | 0.80 (0.49–1.29) | 0.98 (0.72–1.34) | 0.85 (0.58–1.24) | 0.68 (0.41–1.10) |
| Rural (vs urban) | 0.73 (0.44–1.21) | 0.47 (0.17–1.30) | 0.71 (0.38–1.30) | 0.36 (0.15–0.88)* | 0.66 (0.30–1.45) |
| Oromia (vs Addis Ababa) | 3.38 (1.46–7.80)** | 0.97 (0.36–2.64) | 4.96 (1.67–14.69)** | 1.82 (0.70–4.74) | 0.27 (0.08–0.88)* |
| Gambella (vs Addis Ababa) | 1.76 (0.72–4.34) | 0.70 (0.21–2.37) | 2.05 (0.65–6.47) | 1.90 (0.69–5.21) | 1.11 (0.36–3.41) |
| Mother primary education (vs none) | 1.28 (0.74–2.23) | 0.70 (0.33–1.47) | 1.22 (0.64–2.31) | 1.44 (0.90–2.29) | 1.15 (0.65–2.04) |

Population-averaged odds ratios are expected to be closer to 1 than the conditional odds ratios from multilevel models when random-effect variances are large.

**Additional file 1, Table S3.** Interviewers flagged for excess reports of daily anxiety or depression (cluster-stratified Mantel–Haenszel test, Bonferroni p < 0.05/163)

| **Interviewer ID<br>(v028)** | **Region** | **Children<br>(n)** | **Daily<br>anxiety/depression<br>(%)** | **Expected<br>cases<sup>a</sup>** | **Observed cases** | **z** |
|---|---|---|---|---|---|---|
| 1406 | Oromia | 83 | 71.1 | 20.3 | 59 | 12.5 |
| 1508 | Oromia | 97 | 46.4 | 23.6 | 45 | 7.5 |
| 1311 | Oromia | 35 | 62.9 | 8.4 | 22 | 6.6 |
| 406 | Afar | 75 | 21.3 | 5.4 | 16 | 5.7 |
| 1802 | Somali | 20 | 65.0 | 5.0 | 13 | 5.6 |
| 2806 | Gambella | 29 | 27.6 | 2.2 | 8 | 5.1 |
| 1207 | South Ethiopia | 114 | 23.7 | 16.0 | 27 | 4.4 |
| 307 | Tigray | 85 | 11.8 | 4.1 | 10 | 3.9 |
| 3102 | Harari | 17 | 35.3 | 1.8 | 6 | 3.8 |
| 1507 | Oromia | 21 | 42.9 | 3.6 | 9 | 3.7 |
| 706 | Amhara | 79 | 5.1 | 1.1 | 4 | 3.6 |

<sup>a</sup> Expected from the proportion recorded by other interviewers in the same clusters.

